# Genetic diversity of dengue virus serotype 1 associated with rare dengue ophthalmic syndrome in Reunion Island, Southwestern Indian Ocean, 2020-2022

**DOI:** 10.64898/2026.02.15.26346346

**Authors:** Toscane Fourié, David A. Wilkinson, Dana Al Halabi, Jean-Jacques Hoarau, Xavier Deparis, Antoine Bertolotti, Patrick Mavingui

## Abstract

In the past decade, dengue fever has emerged as a major public health on Réunion Island in the Southwest Indian Ocean. During the 2018-2022 outbreak, an unusual increase in ocular complications was reported in some patients. To investigate a potential viral cause, we analysed 447 blood samples from hospitalized patients with and without ophthalmic symptoms. Genetic sequencing revealed the co-circulation of two strains of dengue virus serotype 1, both genetically linked to strains previously identified in Asia. Notably, all patients with ophthalmic symptoms were infected with viruses from a single cluster within genotype I, which harbored several unique mutations. These findings suggest that the rare ocular complications observed during this outbreak may be associated with specific viral cluster. Further laboratory studies are required to confirm this potential link.

## Introduction

Dengue fever is a mosquito-borne disease caused by dengue viruses (DENV), transmitted to humans primarily by *Aedes* mosquito vectors. While traditionally confined to tropical region, the spread of these mosquito vectors into temperate regions has led to the global expansion of the disease [1]. Most dengue infections are mild or asymptomatic, but some progress to severe forms such as dengue hemorrhagic fever or dengue shock syndrome, resulting in around 36,000 deaths annually [2]. Dengue viruses are 11-kilobase, positive-sense, single-stranded RNA viruses in the Orthoflavivirus genus, comprising four antigenically distinct serotypes (DENV-1 to DENV-4), sharing 65% nucleotide identity. Phylogenetic analyses reveal further genetic diversity within serotype [3].

In the Indian Ocean, DENV has been documented since the late 1970’s, notably with DENV-2 outbreak [4–7]. A 1980s survey in Madagascar suggested all four DENV serotypes were circulating [8]. Between 1980 and 2010, localized outbreaks primarily involved DENV-1 and DENV-2, with occasional DENV-3 detection [9]. The past decade experienced DENV re-emergence, including significant outbreaks in Mayotte in 2014 [10], Seychelles in 2016-2017 [11], and Reunion Island (2018-2022), where sustained transmission suggests a shift toward endemicity [9].

Reunion Island’s 2018-2022 outbreak was marked by high incidence peaking at 3,417 cases per 100,000 inhabitants in 2021. About 147,686 cases were reported (48% laboratory-confirmed), with 2,702 hospitalizations and 75 deaths [9,12]. Initially dominated by DENV-2 in 2018-2019 [13,14], the outbreak saw DENV-1 emergence by late 2019 and become the sole circulating serotype by 2020 [15], persisting through 2022 [16]. The outbreak peaked in 2021 and declined to inter-epidemic levels by 2023.

Between 2020 and 2021, Reunion clinicians reported over 100 dengue patients with atypical ophthalmic symptoms, including scotoma (71.4%), sudden decrease in visual acuity (39.2%), with macular involvement in 64.2% [17]. Though are, such ocular complications have been reported globally, mainly in Southeast Asia and the Indian Subcontinent, typically involving retinal vasculitis, maculopathy, or optic neuropathy. Outcomes are often favorable, but permanent impairment has occurred [18,19]. Due to underreporting, estimated incidence ranges from 10 to 40% [20,21]. Despite evidence linking viral mutations to tissue tropism and virulence [22–24], viral determinants in ocular involvement are rarely examined. Of 40 reviewed studies, only five identified serotypes and none included genotyping or full genome sequencing [25–29].

In early 2020, when both DENV-1 and DENV-2 co-circulated in Réunion, preliminary data linked all ocular cases exclusively to DENV-1 [17,30]. The aim of the present study is to explore the genetic diversity of DENV-1 strains circulating during the outbreak in Réunion Island and to examine their potential link to the observed ophthalmic manifestations of dengue infection. To this end, we performed whole-genome sequencing of dengue viruses using blood samples collected in hospitalized patients presenting ophthalmic and non-ophthalmic forms of DENV infections. This approach allowed us to generate 23 full DENV-1 genomes and explore possible viral genetic determinants linked to dengue-associated ophthalmic manifestations.

## Materials and Methods

### Ethics statement

Human blood samples processed anonymously were part of the CARBO (Cohort of patients infected by an ARBOvirus) collection, with ethical clearance granted by the French National Agency for Safety of Medicines and Health Products (ANSM; n°IDRCB 2010-A00282-37) and the committee for the protection of individuals (CPP Sud-Ouest and Outre-Mer III, 06/30/2010), according to French law. Patients provided written informed consent before enrollment.

### Human Sample Collection and Processing

During the DENV 2018-2019 outbreaks in Reunion Island, a joint medical-academic initiative established the prospective observational CARBO cohort, based on a protocol first implemented in the French West Indies during the 2009 outbreak [31,32]. Enrollment and sample collection began in 2019 and continue at the University Hospital of Reunion Island emergency departments. Methods and preliminary findings from the first year were previously published [33]. Briefly, patients presenting within a week of symptom onset and showing at least two dengue-like symptoms were recruited after confirmation. Blood samples were collected at inclusion, and sociodemographic and clinical data were recorded using standardized questionnaires. Follow-ups occurred during convalescent (up to 6 months), with additional blood sampling. Cases were classified by severity following the 2009 WHO dengue guidelines. Ophthalmic manifestations included decrease of visual acuity, scotoma, macular damage or oedema, foveolitis, papilla damage, uveitis, retinal thickening and reflectivity (OCT), and hemorrhages or cotton wool spots (fundus exam) [17].

All biological samples were aliquoted and stored at −80°C by the Centre for Biological Resources of University Hospital. Inclusion-time sera or plasma were sent to UMR PIMIT for genome amplification and sequencing, followed by phylogenetic and mutation analysis, using protocols described by Hafsia *et al* [15].

### Molecular diagnosis and serotyping

Viremia enrolment samples were confirmed by RT-qPCR. Viral RNA was extracted from thawed sera or plasma using the QIAamp 96 Virus Kit and QIAcube HT (Qiagen), according to manufacturer’s recommendations. cDNA synthesis was performed with Lunascript™ RT SuperMix (NEB) using random hexamers and 10 µM DENV-specific reverse primers (Den123_R10735, Den4_R10653), as published [34], in a final 12µL reaction, then stored at - 20°C. Samples were screened with a pan-DENV qPCR system; positive samples were serotyped using DENV-specific qPCRs [35]. All qPCRs were run on the CFX96™ system (Bio-Rad) using QuantiNova Probe qPCR kit (Qiagen).

### Genome sequencing

cDNA of DENV-1 positive samples (ct<34) was used for full-genome amplification via an amplicon tiling protocol targeting four overlapping genomic regions [34]. Substitution primers were used to fill amplification gaps and improve complete coding sequence (CDS) recovery [36]. For samples where full-genome amplification failed, targeted primers were used to amplify the envelope (E) gene, a reliable DENV genotyping marker [3,37]. As the first ∼ 2500 nt fragment includes the E gene, its amplification (via three overlapping fragments) was only attempted when fragment 1 failed [36]. PCRs used Q5 Hot-start High-Fidelity DNA Polymerase (NEB), and amplification was confirmed by gel electrophoresis. Samples with complete overlapping fragments covering or including the full E-gene were selected for sequencing.

Sequencing followed Oxford Nanopore Technologies Native Barcoding protocol (EXP-NBD104) on the Mk1C device with R9.4.1 flowcells [38] [15,39]. Briefly, equimolecular PCR products were dA-tailed using the NEBNext® Ultra™ II End Repair/dA-Tailing Modul, barcoded via blunt-end ligation, and purified with AMPure-XP SPRI (Beckman Coulter). Sequencing followed manufacturer protocols and stopped after achieving 1000x predicted coverage. Basecalling and demultiplexing were done with Guppy v.6.5.7 using the super-accurate model v4.3.0. Cutadapt v5.0 trimmed primers, and reads were mapped to reference sequence GCF_000862125.1 using minimap2 v2.25. Samtools v1.21 generated consensus sequences, followed by error-correction with medaka v1.12.0. Genomes with >5% missing data or a median depth < 300× were excluded. All complete CDS and E-gene sequences obtained were deposited in GenBank under accession numbers [PV687461 to PV687470; PV687481; PV687483].

### Phylogenetic analysis

Phylogenetic analyses were performed DENV-1 CDS and E-gene sequences. Alignments and maximum-likelihood trees followed the approach in Fourié *et al* [40]. The CDS dataset was adapted by adding all DENV-1 sequences from SWIO islands or with 99% identity to study sequences. The E gene dataset included all CDS plus GenBank sequences meeting the same geographic and identity criteria. Datasets was curated by removing incomplete, ambiguous, redundant, metadata-deficient sequences as per [40], resulting in 142 complete CDS and 154 E-gene sequences (<u>see supplemental tables S1 and S2</u>). Sequences were aligned with MAFFT (v7.511) [41] and trimmed. Maximum-likelihood trees were built with IQ-Tree (Galaxy platform) using GTR+F+I+G4 (BIC, –ufboot 1000, –bnni) [41–44]). Final trees were midpoint-rooted and annotated following Weaver and Vasilakis classification of 2009 [3], using iTol v6.9.

### Mutation analysis of the Genotype 1 Reunion Island 2019-2022 outbreak group

To examined amino acid (AA) substitutions linked to ophthalmic cases, all available DENV-1 genomes from Reunion (n=555) were retrieved from GenBank. Sequences >9000 nt and <50 ambiguous nucleotides were included. AA alignments were generated by integrating new sequences into the CDS and E-gene datasets, followed by alignment, translation, and trimming to conserved regions. Hierarchical Bayesian Population Structure analysis (hierBAPS, rhierbaps v1.1.4) clustered sequences by nucleotide similarity.

To assess clustering of ophthalmic cases, maximum likelihood phylogenies were built from CDS and E gene alignments. The most recent common ancestor (MRCA) node encompassing all ophthalmic cases was identified in the E-gene tree, which captures the largest number of such cases. This cluster was then mapped onto the full-genome phylogenetic tree, defining the associated group of strains as the “ophthalmic ingroup”.

For mutation analysis, the ophthalmic ingroup was compared to the “outgroup”, consisting of all other sequences in the complete CDS. Two additional outgroups were defined for resolution: “Outgroup 1” included closely related genotype 1 sequences (as per rhierbaps), and “Outgroup 2” included more distantly related genotype 1 sequences. These groups are illustrated in Figure 1. AA variation was assessed across 3367 aligned protein positions (excluding the final 25 NS5 N-terminal positions). The analysis focused on conserved mutations within the ingroup, defined as AAs present in ≥95% of the ingroup sequences and absent or infrequent (< 50% of sequences) in the outgroup.

**Figure 1.**
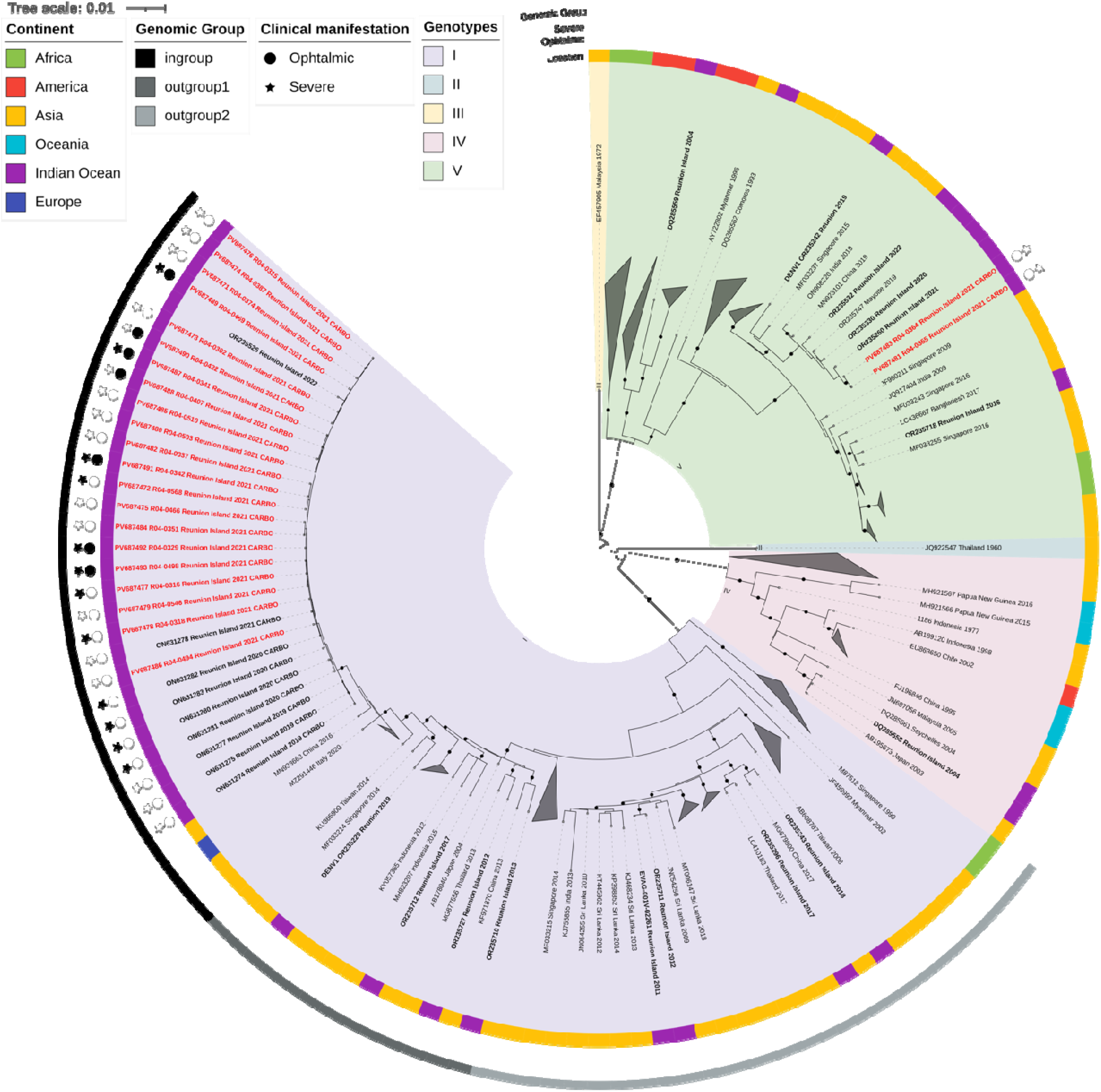
Maximum-likelihood phylogenetic tree based on 172 complete CDS of DENV-1 sequences (9491 nucleotide-long sequences after trimming). DENV-1 genome sequences from Reunion Island are indicated in bold and those from the CARBO study are red lettering (including 8 previously published strains). Each sequence is identified by its GenBank accession number, country of origin and year of sample collection. The tree was rooted at the mid-point and bootstrap values equal to 100% are indicated by a black circle on branch. Scale bar indicates genetic distance (nucleotide substitutions per site). Genotypes are represented with colored overlay; color strip represents location of infection; clinical manifestations of cases in the CARBO cohort are represented with stars for severe cases and circle for ophthalmic cases. DENV, indicates dengue virus.

## Results

### Study population and sample overview

Between 2019 and 2021, 447 patients were enrolled in the CARBO study, including 81 patients with ophthalmic manifestations. Blood samples were collected on average 6.64 days (SD: 3.49) after symptom onset, 9.11 days (SD: 4.36) for ophthalmic cases and 5.61 days (SD: 2.40) for non-ophthalmic cases. A subset of the 129 cases met the criteria for molecular investigation. Among them, 121 collected in 2021 were eligible for genome amplification (44 serum and 77 plasma samples). Additionally, eight sequences from 2019-2020 CARBO cases [15] were included (GeneBank ascension numbers: ON631274, ON631275, ON631277, ON631278 and ON631280 to ON631283). Among 129, 43 (35%) had ophthalmic symptoms, and 84 (n=36) were severe dengue.

### Genomic and phylogenetic analyses

DENV RNA was detected in 54% of blood samples collected during the early stages of enrolment in the 2021 cohort (n=65), with detection up to 11 days after symptom onset. Among these, 88% were identified as DENV-1 (n=57), while the remaining samples could not be serotyped, mostly due to late inclusion after symptom onset with an average of 9 days (range 6 to 11 days). No cases of DENV-2, DENV-3, or DENV-4 were detected. Of the 57 samples with positive RT-PCR, genome amplification was attempted for 44, resulting in 23 complete CDS and 10 additional E-gene sequences. Genomic data were also successfully obtained from 29% of ophthalmic cases (n=12) from the 2021 collection.

Phylogenetic tree reconstructions based on both complete CDS and the E-gene sequences revealed that the DENV-1 sequences from our sample set belonged to two distinct genotypes, unevenly represented (Figure 1 and 2). Two of the 34 sequences from 2021 Reunionese CARBO cases clustered within the Asian lineage of genotype V, closely related to isolates previously detected in Reunion Island in 2020 (OR235330) and Mayotte Island (French Overseas territory of the Comoro archipelago, Indian Ocean) in 2019 (OR235747), sharing 99.7% nucleotide identity (Figure 1). Phylogenetic analysis of E-gene showed close relationships to sequences reported in China (MN923101), India (ON908220) and Tanzania (LC485151) in 2019 (Figure 2). Based on complete CDS phylogeny, this genotype V cluster is distinct from earlier Reunion strains of the same lineage identified in 2016 (OR235718) and 2019 (OR235242), suggesting distinct separate introduction events (Figure 1).

**Figure 2.**
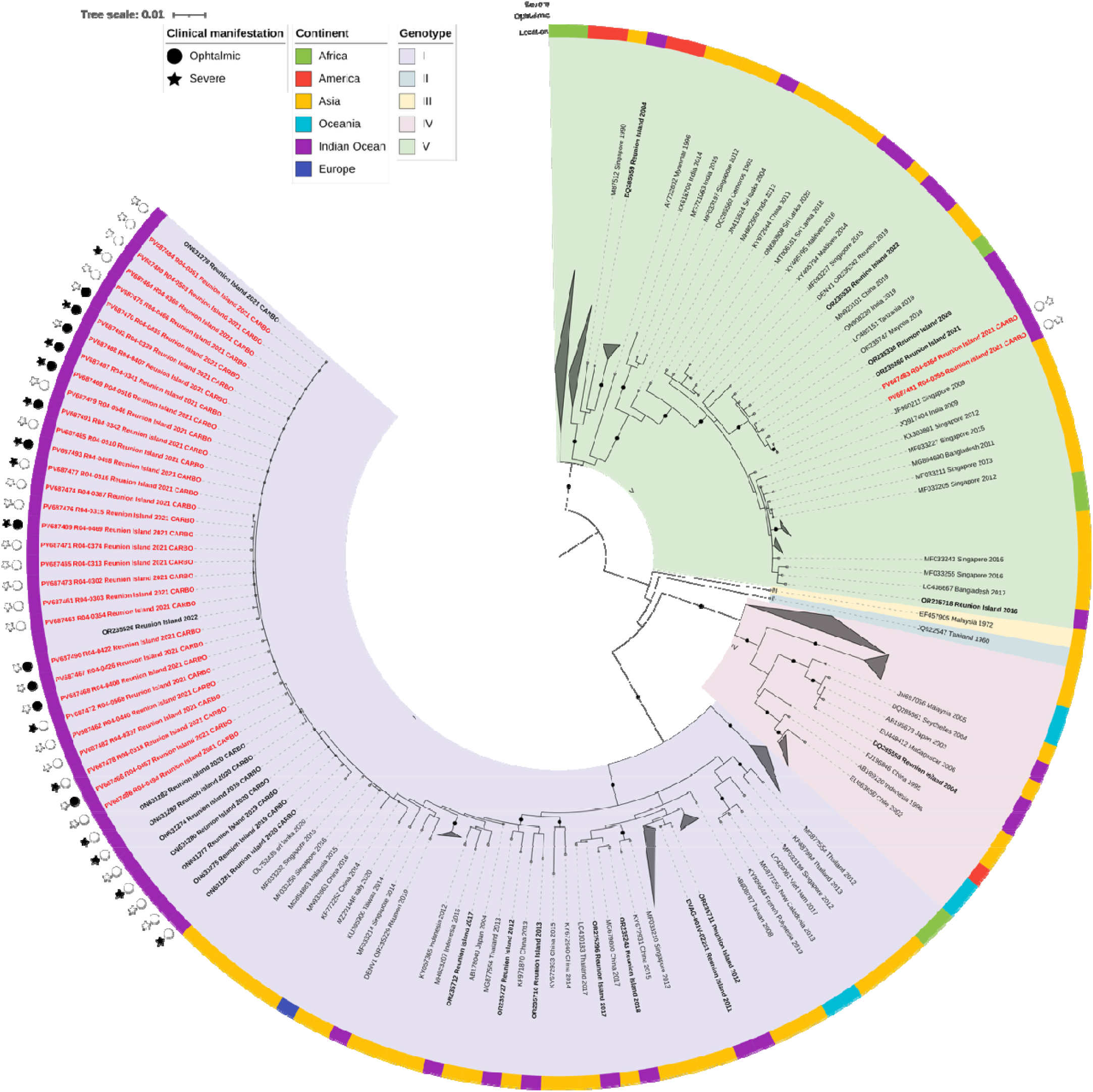
Maximum-likelihood phylogenetic tree based on 195 complete E-gene of DENV-1 sequences (1485 nucleotide-long sequences after trimming). DENV-1 sequences from Reunion Island are indicated in bold and sequences from the CARBO study are red lettering (including 8 previously published). Each sequence is identified by its GenBank accession number, country of origin and year of sample collection. The tree was rooted at the mid-point and bootstrap values equal to 100% are indicated by a black circle on branch. Scale bar indicates genetic distance (nucleotide substitutions per site). Genotypes are represented with colored overlay; color strip represents location of infection; clinical manifestations of cases in the CARBO cohort are represented with stars for severe cases and circle for ophthalmic cases. DENV, indicates dengue virus.

The remaining 32 sequences from 2021 fell within genotype I, along with seven 2019-2020 CARBO sequences and 435 additional Reunion sequences (2019-2022) published by the local reference center (represented by sequence OR235526 on phylogenetic trees. This genotype I cluster, hereafter referred to as the Reunion Island 2019-2022 outbreak group, shares more than 99.7% nucleotide and AA identity. Phylogenetic analyses of both complete CDS and E-gene showed the closest known relatives of this group to be strains from continental Asia (MN933663) and Sri Lanka (OL752439) (Figure 1 and Figure 2). Importantly, this Reunion Island 2019-2022 outbreak group is phylogenetically distinct from other genotype I strains sporadically detected in Reunion Island since 2011 and 2019 (Figure 1). The complete CDS maximum likelihood tree indicates the presence in Reunion Island of 8 distinct strains of DENV-1 genotype I, primarily known to be circulating in Asia.

Regarding clinical manifestations, all severe and ophthalmic cases were associated with the Reunion Island outbreak group genotype I. In contrast, the two genotype V stains were retrieved from both non-severe and non-ophthalmic cases.

### Mutations harbored by the Genotype 1 Reunion Island 2019-2022 outbreak group

After excluding incomplete sequences, 506 La Reunion-associated sequences were analyzed. The majority of these sequences originated from a single data submission [16]. Phylogenetic clustering showed that sequences contemporary to the 2019–2022 outbreak period grouped closely with those from our clinical cohort. Specifically, most (n=455) fell within the 2019-2022 genotype I, while 16 additional sequences clustered with the two genotype V sequences identified in our dataset.

To assess links to ophthalmic cases, the MRCA node from the CDS phylogenetic tree was examined. This node encompassed the entire outbreak group and five older related strains detected prior to the outbreak: KP772252 and MN933663 from China (2014 and 2016), MF033250 and MF033232 from Singapore (2015 and 2016), and MZ291446 from Italy (2020) (Figure 1, Figure 2). These findings indicate that ophthalmic cases were not confined to a unique phylogenetic subcluster, but rather distributed throughout this broader lineage.

Consequently, the ophthalmic ingroup comprises to full 2019-2022 outbreak cluster and the five older, related strains, totaling 486 sequences (Figure 1). These sequences share 98,9% AA identity across 3164 AA positions. When the 5 older strains were excluded, AA identity within the outbreak group rose to 99,99%, highlighting its genetic homogeneity. Three comparison groups designated “outgroup”, “outgroup 1” and “outgroup 2”, included 198, 26 and 36 sequences, respectively. In total, 11 AA mutations fitting defined selection criteria were identified, all found in ophtalmic cases sequences. All mutations occurred in non structural proteins, with six located in the NS5 protein. Of these, nine mutations were unique to the ophthalmic ingroup and absent from both outgroups 1 and 2. Notably, the NS5:S566A mutation appeared in both the ingroup and most of outgroup 1 but was absent from outgroup 2. Conversely, the NS3:E12K mutation was shared between outgroup 2 and the 2019-2022 outbreak group, while outgroup 1 retained the DENV-1 consensus AA. Most mutations were conservative (*i*.*e*. involving substitution of amino acid with similar biochemical properties). The exception was NS5:S566A, located within the RNA-dependent RNA polymerase (RdRp) domain, representing a non-conservative change from polar serine to non polar alanine.

## Discussion

### Genetic heterogeneity of DENV-1 outbreaks in Reunion Island

Our phylogenetic analysis confirms that the 2019-2022 dengue outbreak in Reunion Island was predominantly driven by a DENV-1 genotype I strain, with a limited co-circulation of a genotype V [15,16]. Beyond this, our data highlight the notable genetic heterogeneity of DENV-1 in Reunion over the past two decades. Prior to 2019, the island experienced multiple introductions of genetically distinct strains, typically resulting in small outbreaks, localized clusters, or sporadic cases, including those diagnosed in travelers to mainland France [9].

Most introductions were linked to viral lineages circulating in Asia, with spillover into the Indian Ocean, Africa, and Europe. At least 12 events were traced to Southeast Asia, the Indian subcontinent, or China; eight involving genotype I introduced since 2011. Three sequences were from travelers returning to Europe (strains DQ285558, DQ285559 and EVAG-CNR14446). E gene phylogeny further illustrates DENV’s global dispersal, connecting Asia to East Africa and SWOI islands (*e*.*g*., 2019-2020 genotype V cluster), as well as inter-island transmission within SWOI (*e*.*g*., 2004-2006 genotype IV cluster in Reunion, Madagascar and Seychelles).

While phylogenetic analysis is subject to sampling biases, increasing sequence data is enhancing our understanding of global DENV circulation. Reunion’s strategic position—with cultural, commercial, and travel ties with Asia, Africa, and Europe—amplifies its vulnerability to introductions. The island’s links to dengue-endemic regions (e.g., Thailand, India, Singapore) and over 600 annual international cargo/cruise ship stopovers (2020 data) increase the risk of DENV import/export via infected travelers or mosquitoes.

### Emergence of ophthalmic manifestations during the 2019-2022 outbreak

Ocular involvement in dengue fever has been sporadically reported since the 1970’s, with increased frequency after 2004, notably in Singapore and India [45]. In Singapore, ophthalmic symptoms occurred in ∼5-6% of dengue cases, particularly during DENV-1 outbreaks [45-47]. However, no genetic studies of ophthalmic-associated DENV-1 strains from Singapore are publicly available.

In Reunion Island, ocular cases were first reported in 2020, following the replacement of DENV-2 by DENV-1. These cases peaked during the 2019 to 2022 outbreak. Phylogenetic analysis revealed uneven co-circulation of two Asian-lineage DENV-1 genotypes. Our genomic investigation detected only genotype I in viremic ophthalmic cases from 2020-2021, while two individuals infected with genotype V showed no ocular manifestations. All sequenced ophthalmic cases were infected by the predominantly circulating genotype I strain. However, the low number of genotype V infections limited the statistical power to evaluate this association.

A key limitation is that the 2020 expansion of the inclusion criteria beyond the first week of symptom onset reduced the likelihood of sampling during viremia. Since DENV typically clears from bloodstream within 7-10 days, this likely explains the low RNA rate after day seven. Despite this, our study provides detailed molecular data linking specific DENV strains to ophthalmic manifestations, supporting a possible association genotype I.

### Potential viral genetic determinants linked to ophthalmic complications

Genomic analysis showed that strains from ophthalmic cases were distributed throughout the genotype I cluster, indicating that any strain within this lineage could be implicated. We identified 11 unique mutations in DENV-associated ophthalmic cases, all in nonstructural proteins: two in NS1, two in NS2B, one in NS3 and six in NS5. The NS5 changes were in the RNA-dependent RNA polymerase (RdRp) region. While most mutations were conservative, the S566A substitution (serine to alanine) in RdRp was non-conservative and may impact enzymatic function, replication fidelity, or host interaction [47]. The consistent presence of these mutations, especially in strategic protein domains, suggests potential roles in enhanced pathogenicity or ocular tropism. Functional studies are needed to explore these hypotheses.

### Other determinants and perspectives

Given that the DENV-1 outbreak followed the DENV-2 wave, the risk of severe disease due to secondary infection (via antibody-dependent enhancement) may have been elevated [48]. Therefore, assessing the serological status of ophthalmic patients is crucial. Moreover, the timing of ocular symptoms, typically during early platelet recovery, supports the role of immune-mediated mechanisms [18,49]. Clinical outcomes likely result from complex interactions among host immunity, viral genetics, and host genomics. Compared to viruses like SARS-CoV-2 [50], the genetic basis for clinical variability in dengue remains poorly defined. Nevertheless, our findings underline the importance of considering viral genetics in atypical dengue presentations. We encourage ophthalmologists and researchers to continue reporting publishing molecular data from dengue-associated ocular cases.

## Conclusions

This study provides a detailed molecular characterization of DENV-1 strains from hospitalized patients during the 2019–2022 outbreak in Réunion Island, with a focus on cases presenting with ophthalmic complications. Phylogenetic analysis confirmed the predominance of genotype I, with occasional genotype V circulation, reflecting broader population trends. All ophthalmic cases were linked to the Reunionese cluster within genotype I. Genomic analysis did not identify a unique molecular signature specific to ophthalmic cases. However, the Reunion 2019-2022 cluster showed high AA homogeneity (99,99%) and shared 11 conserved mutations in non-structural proteins, several within the NS5 RNA-dependent RNA polymerase domain. While most substitutions were conservative, their combined presence, particularly the non-conservative change S566A, may suggest an impact on viral fitness or tissue tropism.

Although our findings suggest a possible link between viral genotypes and atypical clinical outcomes, the evidence is preliminary due to small sample size and lack of functional validation. Further research integrating host immune profiling and experimental virology is needed to clarify the mechanisms underlying dengue-associated ocular disease and broader pathogenic variability.

## Data Availability

All data produced in the present work are contained in the manuscript

## Acknowledgments

The authors thank Daed El Safadi, Mathieu Pot, Loic Raffray, Quentin Richier, Loraine Gauzere, Eric Doussiet, Patrick Gerardin, Lucas Balloy, Stella Hoang, Cecile Saint Pastou, Marie-Pierre Moiton, Fréderic Renou, Olivier Maillard, and Yves-Marie Diarra for their support in epidemiological and laboratory investigations.

---

**Table 1.** Demographic and clinical characteristics of our cohort, Reunion Island, 2019–2021.

|  | *Non-ophthalmic cases*<br>*(n=86)* | *Ophthalmic cases*<br>*(n=43)* | *Total cohort*<br>*(n=129)* |
| --- | --- | --- | --- |
| ***Sampling year*** |  |  |  |
| 2019 | 4 (4.6%) | - | 4 (3.1%) |
| 2020 | 4 (4.6%) | - | 4 (3.1%) |
| 2021 | 78 (90.8%) | 43 (100%) | 121 (93.8%) |
| ***Sex*** |  |  |  |
| female | 41 (47.7%) | 26 (60.5%) | 67 (51.9%) |
| male | 45 (53.3%) | 17 (39.5%) | 62 (48.1%) |
| ***Age (years)*** |  |  |  |
| 15–24 | 8 (9.3%) | 6 (9.3%) | 14 (9.3%) |
| 25–34 | 7 (2.3%) | 14 (14.0%) | 21 (6.2%) |
| 35–44 | 14 (11.6%) | 10 (14.0%) | 24 (12.4%) |
| 45–54 | 16 (10.5%) | 7 (4.7%) | 23 (8.5%) |
| 55–64 | 14 (32.6%) | 4 (30.2%) | 18 (31.8%) |
| 65–74 | 18 (27.9%) | 2 (23.3%) | 20 (26.4%) |
| 75+ | 8 (8.1%) | - | 8 (0.8%) |
| median (range) | 54 (15-84) | 35 (16-74) | 46 (15-84) |
| ***Sample type*** |  |  |  |
| plasma | 60 (69.8%) | 25 (58.1%) | 85 (65.9%) |
| serum | 26 (30.2%) | 18 (41.9%) | 44 (34.1%) |
| ***Severe clinical form*** |  |  |  |
| severe | 26 (30.2%) | 36 (83.7%) | 62 (48.1%) |
| non-severe | 60 (69.8%) | 7 (16.3%) | 67 (51.9%) |

**Table 2.** Mutations identified in the genotype I Reunion Island 2020–2022 outbreak group (also defined as ingroup of ophthalmic cases) and their distribution among genomic groups.

| Protein | Mutation Name | Mutation type | Consensus AA within ingroup | Frequency within ingroup | Frequency within outgroup | Consensus AA in outgroup1 (frequency) | Consensus AA in outgroup 2 (frequency) |
| --- | --- | --- | --- | --- | --- | --- | --- |
| **NS1** | R214K | Conservative | K | 100% | 38% | R (100%) | R (100%) |
|  | N293S | Conservative | S | 100% | 38% | N (100%) | N (100%) |
| **NS2B** | I44M | Conservative | M | 100% | 36% | I (100%) | I (100%) |
|  | V106I | Conservative | I | 100% | 47% | V (100%) | V (100%) |
| **NS3** | E12K | Conservative | K | 100% | 44% | E (96%) | K (67%) |
| **NS5** | K388R | Conservative | R | 100% | 36% | K (100%) | K (100%) |
|  | V413I | Conservative | I | 100% | 41% | V (64%) | V (100%) |
|  | K551R | Conservative | R | 95% | 11% | K (100%) | K (82%) |
|  | E559D | Conservative | D | 100% | 36% | E (96%) | E (100%) |
|  | S566A | Non-conservative | A | 100% | 46% | A (88%) | S (100%) |
|  | V686I | Conservative | I | 99% | 36% | V (100%) | V (100%) |

